# Immune Reconstitution Inflammation Syndrome Incidence, Demographic Indicators, Survival-Time to and Prediction of Adverse Pregnancy Outcomes: A Kaplan Meier and Cox-Regression Analysis

**DOI:** 10.1101/2024.11.28.24318132

**Authors:** John Kyalo Muthuka, Lucy Kathure Manyara, Agnes Muthoni Linus, Kelly Oluoch Jack, Lucy Chepkemei Chebungei, Rosemary Nabaweesi

## Abstract

**Background and aim:** There are persistent concerns regarding the potential adverse effects of *in utero* ART exposure. Whereas the association between untreated, advanced HIV disease and adverse pregnancy (APOs) outcomes is well documented, non or few have focused on APOs subject to immune reconstitution inflammatory syndrome (IRIS) as a predictor, devoid of differential aspect of either paradoxical or unmasking IRIS. The current study sought to investigate the incidence and demographic indicators of IRIS, IRIS type, survival-time to, and its prediction of APOs.

**Methods:** An active records study was conducted between June 2019 and March 2020 among ART-naïve pregnant women attending the antenatal care units (ANCu) at the Kenyatta National and Mbagathi Hospitals, Nairobi, Kenya. Participants were aged between 20 and 49 years and had a confirmed HIV-positive test. IRIS diagnosis was adjudicated for accuracy and consistency by an independent review committee. Baseline demographic characteristics including: age, education level, religion, marital status, residence, occupation and economic status were recorded. IRIS incidence was assessed using the International Network for Studies Against HIV-Associated IRIS (INSHI) during the first three months after ART initiation. Bivariate analysis via Pearson’s test for demographics relative to IRIS type was performed. The association and its strength between the IRIS type and APOs were further established through Pearson Chi-Square test and Phi and Cramer’s V tests respectively. Kaplan Meier analysis test estimated the survival time to APOs using log rank test statistic. Multivariate Cox-regression analysis for Pre-ART demographics on IRIS type incidence was performed using survival package in SPSS. Decision tree analysis for predictive modelling on APOs relative to demographics was conducted.

**Results:** The incidence of IRIS was 25% (n=133) among the 532 ART-naïve pregnant women with 97 (72.9%) presenting with unmasking IRIS, significantly associated with APOs [χ (1) = 4.911^a^, *P* = 0 .027]. Maternal age, between 40-49 years had a positive co-efficient with unmasking IRIS [(β)=0.329, Wald test (β^) = 1.011, (HR = 1.389, 95% C.I 0.732 - 2.638, P = 0.325]. The cumulative survival function evaluating all the demographic characteristics (as covariates) indicated that, over 80% of the ART naïve pregnant women survived IRIS diagnosis for six weeks, while half of them had been diagnosed with it at approximately two months. Kaplan-Meier survival function showed that, women diagnosed with unmasking IRIS compared to those diagnosed with paradoxical IRIS survived longer before experiencing an APO (*χ2 =* 5.292, Log Rank test = 0.021), cumulative hazard, [HR = 0.18 and 0.4] respectively. Decision tree analysis demonstrated that, women aged 30-39 had most of APOs (P = 0.688).

**Conclusion:** Unmasking IRIS was the most common, associated highly with APOs that were experienced much later compared to those predicted by paradoxical IRIS with older age being a plausible predictor. The survival time to experiencing an APO was longer for women presenting with unmasking IRIS as opposed to paradoxical IRIS, supporting the need to conduct further research on possible APOs due to paradoxical IRIS.

## Background Information

The immune recovery associated with ART is crucial in pregnant women but can be complicated by IRIS with deterioration of clinical manifestations that follows successful suppression of HIV viremia with ART [1]. Early initiation of ART is recommended in patients with CD4 <50 cells/µL before opportunistic infections are adequately treated[2].

There are benefits of ART for maternal health and prevention of perinatal transmission, however, some risks are or may be possible with specific type of IRIS [3]. The key features of IRIS include clinical deterioration in the first weeks to months of ART, with evidence of localized tissue inflammation with or without a systemic inflammatory response[4], [5].There are persistent concerns regarding the potential adverse effects of *in utero* ART exposure.

Whereas the association between untreated, advanced HIV disease and adverse birth outcomes is well documented [6], a number of studies have suggested increased levels of preterm (PTD)[7], low birthweight (LBW), and/or small for gestational age (SGA) deliveries among women receiving ART[3], [8], [9], [10], [11].

In addition, most prospective studies on IRIS in pregnancy have focused on APOs in relation to IRIS as a predictor without ascertaining either paradoxical or unmasking. In this regard, it is not clear to what extent these relationships apply by the type IRIS, where overall findings for the putative association between antenatal ART use which is linked to IRIS and APOs are highly mixed[8], [12], [13], [14]. These APOs are also associated with other HIV-related factors, including progression stages of HIV, antiretroviral therapy, CD4 cell count, viral load and injection drug use[15], [16].

Given the large numbers of ART-exposed pregnancies around the world[17] and the conflicting evidence to date, better understandings of the potential associations between the specific type of IRIS and APOs as well as estimating hazard and survival rates to experiencing an APO are critical. Further, some key existing trial report has shown that, use of the most common combination antiretroviral medicines in pregnancy have been questioned [18].

This research was conducted to estimate the IRIS cumulative incidence and the associations with APOs. Further, it determines baseline demographics as predictors of, and analyzes survival time to an APO relative to specific type of IRIS by such predictors among ART-naïve pregnant women visiting a maternal and child healthcare clinic (MCH) for the first time and put on ART in the first trimester before assessing the specific type of IRIS outcome. A decision tree analysis was conducted to establish any possible specific complex interactions between baseline demographics and APOs.

## Materials and Methods

### Study overview

The current hospital-based, active records study was conducted among ART-naïve, HIV-positive pregnant women who visited MCHs for the first time during the first trimester before the antenatal care services, and were diagnosed with IRIS (either unmasking or paradoxical) at the Kenyatta National and Mbagathi hospitals in Nairobi, Kenya, from June 1, 2019, to March 30, 2020. The study sought to investigate the incidence and demographic indicators of IRIS, IRIS type and its APOs prediction. Further, it aimed at establishing the rate at which an APOs occurred relative to IRIS and, finally, through decision tree analysis, assess the complex interactions of demographic risk factors for either paradoxical or unmasking IRIS diagnosis.

### Inclusion and exclusion criteria

Pregnant ART-naïve and HIV-positive women, between 20 and 49 years of age, diagnosed with IRIS, in the first trimester, having conceived not later than one month at the first visit (the gestational age based on the date of the last menstrual period (LMP) concept), willing to participate in the study and consented to utilization of their active records were included. Pregnant women who were ART-naïve and HIV-positive, aged below 20 years or above 49 years, with unclear IRIS diagnosis, pregnant for more than one month at the first visit, and with known critical and any severe threatening conditions, were excluded. Participants unwilling to participate in the study at the enrollment stage and declined to give consent for the utilization of their active records were also excluded.

### Sample size calculation

This was determined as per the following formula:

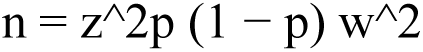

with (n) being the minimum sample size, (w) the estimated error (0.05), (p) the estimated HIV prevalence among pregnant women in Sub-Saharan Africa based on systematic reviews and meta-analysis (9%), and z=1.96 by assuming a 95% confidence interval, 125 IRIS cases would at least be needed. Considering possible attrition, 5% was added, and the total sample was 133.

### Study procedure

Baseline demographic characteristics were recorded among ART-naïve, HIV-positive pregnant women. The cohort of confirmed HIV-positive pregnant women was initiated on ART as a single population at baseline before establishing IRIS diagnosis/outcome. The ART-naïve, HIV-positive pregnant women’s active records from the time they were started on ART to third month (week-12) were reviewed and considered. It is established that the duration of IRIS symptoms typically occurs in two to three months after ART initiation [8]. The ART combination was chosen according to HIV treatment guidelines in pregnancy as guided by the WHO and the clinicians’ case-by-case recommendations.

The clinical teams prospectively identified IRIS events and collected relevant clinical information that was presented to an endpoint review committee comprising an HIV specialist, a nurse specialist, and a maternal health expert and was meant to ascertain and complement through an independent judgment that, the events were consistent with the AIDS Clinical Trials Group definition criteria for IRIS [9], which includes evidence of ART initiation with resultant increase in CD4 count (≥50 cells/µL or a ≥2-fold rise) and/or virologic suppression (>0.5 log10 decrease in plasma HIV viremia), clinical presentation consistent with an infectious or inflammatory condition, and the absence of an alternative etiology such as the expected course of a previously recognized infection or side effects of medications.

### Assessment

#### General baseline demographic and non-demographic characteristics

Basic demographic characteristics – the demographic characteristics of the participants including; age, gender, ethnicity, and EFU composition, number of children (parity), educational attainment, source of income, and socio-economic status were recorded.

#### General Blood Tests

From the potential female population that would produce IRIS cases for the study after ascertaining HIV-positive status, the HIV test (immunoassay for HIV-1 and HIV-2) was done at the same time as other routine antenatal blood tests (blood group and rhesus factor, full blood count, hepatitis B, rubella, and syphilis) were performed.

#### History

A complete medical and family history, past and current comorbidities, any possible concomitant medicines, and psychosocial history of the current lifestyle, was performed during HIV diagnosis prior to ART initiation.

#### Gestational Age Estimation

LMP method assessed the estimated age of the pregnancy to ensure it was not above one month after conception.

#### Co-infections Assessment

Syphilis serology, sexually transmitted infection screening, tuberculosis chest X-ray, and other comorbidities, including SARS-CoV-2, were assessed based on the WHO guidelines among HIV-infected pregnant women.

#### Assessment for IRIS events

The diagnosis of IRIS was examined and confirmed in the first three months after starting ART. This IRIS events were based on INSHI criteria: (1) new or worsening infectious or inflammatory symptoms; (2) >1 log decrease in viral load; and (3) the absence of three other explanations (newly acquired infection, predicted course of previously diagnosed infection, and adverse drug effects). Experts’ opinions further defined the IRIS diagnosis based on signs and symptoms to ensure internal validity.

#### Adverse pregnancy outcome assessments

Any form of adverse pregnancy outcome (APO) from the end of first trimester to post-delivery period was ascertained relative to IRIS type. To substantiate active records information from participants’ files, adoption of three methods, including outpatient follow-up by antenatal care providers (the main method), telephone and home visit (supplementary methods) were done as deemed applicable.

Stillbirth was defined as a fetal death occurring after 20 complete weeks of gestation. Birth < 37 gestational weeks was regarded as PTB, and birth weight <2500 g was considered as LBW. Birth weight <10th percentile was defined as SGA, adjusted for gestational Apgar score referred to the summary of appearance, pulse, grimace, activity and respiration at 1 minute, and neonatal asphyxia was defined as Apgar score ≤7 at 1 minute. Birth defects were classified according to International Classification of Diseases (Tenth Revision).

### Statistical analysis

Descriptive univariate analysis considering IRIS cumulative incidence and by IRIS type was performed. To investigate the relationship between the demographics: age, gender, religion, marital status, number of children (parity), education level, source of income, occupation and socio-economic status with IRIS in the first phase of the study (12 weeks after starting ART), a bivariate analysis was performed using Pearson’s test. To ascertain the association and strength between the primary outcome (IRIS) and the secondary outcome (APOs), further, Pearson Chi-Square test, Phi and Cramer’s V tests. Kaplan Meier analysis test was used to estimate the survival time to an APO experience using log rank test statistic. For the baseline demographic indicators and time to a specific type of IRIS, multivariate Cox regression analysis with IRIS as the outcome was fitted, with censoring effect included prior to 12 weeks post ART initiation, using the *Survival* package in SPSS [19]. To explore further key relationships between baseline characteristics and IRIS-type diagnosis, a decision tree analysis was performed.

### Ethical considerations

Ethical approval was obtained from the University of Nairobi-Kenyatta National Hospital Ethics Committee (ethical approval number: P609/08/2018) before starting the study. Written informed consent to participate and utilize the active records of eligible ART-naïve, HIV-positive pregnant women at the enrollment stage was also obtained. Authority to carry out the research was provided by participating facilities’ research management units (Kenyatta National Hospital, reference number: KNH-ERC/RR 305, and Mbagathi Hospital, reference number: MDH/RS/1/VOL 1). As per the Declaration of Helsinki and local regulatory requirements, the study was conducted in accordance with the principles outlined. Confidentiality and privacy of participants’ information were strictly maintained throughout the study.

## Results

A total of 512 HIV-infected, ART-naive pregnant women were screened; 143 with seemingly signs and symptoms of IRIS identified, of which 133 remained following exclusion of ten (n=10) due to lack of consensus on IRIS presentation at phase one of the study (trimester one) as well as other critical conditions. The 133 were enrolled (91 at Kenyatta National Hospital, and 42 in *Mbagathi* Hospital between August 2019 to May 2020. The majority (53%, n=70) were between the ages of 30-39, 51% (n = 69, with normal body mass index (18.5-24.9), and 80 (60%) were married. Most, 50% (n= 67) had a parity of 2-3. Over 50% (n = 73), were at the first stage of HIV infection as determined in WHO guidelines[20] and 5.3% (n = 7) had hemoglobin level below <11g/dl with 42% (n = 56) presenting with at least a symptom of an opportunistic infection. On ART combination, Tenofovir alafenamide/emtricitabine (TAF/FTC) was started in 72% (n = 96) of the pregnant women initiated on ART while few, 5.3% (n=11) were put on tenofovir disoproxil fumarate/emtricitabine (TDF/FTC; *table 1*.

### Cumulative incidence of IRIS and association with baseline demographics

One hundred and thirty-three, (133) (25%) women experienced IRIS by the 12^th^ week post ART initiation. The proportions of IRIS by type were 27.1 % (n= 36) and 72.9% (n = 97) paradoxical and unmasking respectively. Unmasking IRIS as compared to paradoxical IRIS demonstrated seemingly higher proportions relative to parity distributions, with the highest occurrence being among women who had a parity of one (43 %) (n = 42), very closely followed by those with a parity of 2-3 (42%) (n = 40) with a parity of more than 5 recording unmasking IRIS experience at 14 % (n = 13). On paradoxical IRIS, women with a parity of 2-3 had the highest proportion as compared to other parities (72.2%) (n = 26). Generally, the Pearson Chi-square statistic showed a significant association between parity and IRIS type (χ2 = 10.667^a^, P = 0.014). A positive rhesus factor screening in the first trimester was at a proportion of 91.7 % (n = 88) within IRIS type and significantly associated with unmasking IRIS (χ2 = 4.741^a^, Fisher’s Exact Test P value = 0.034); *table 2*.

**Table 2:**
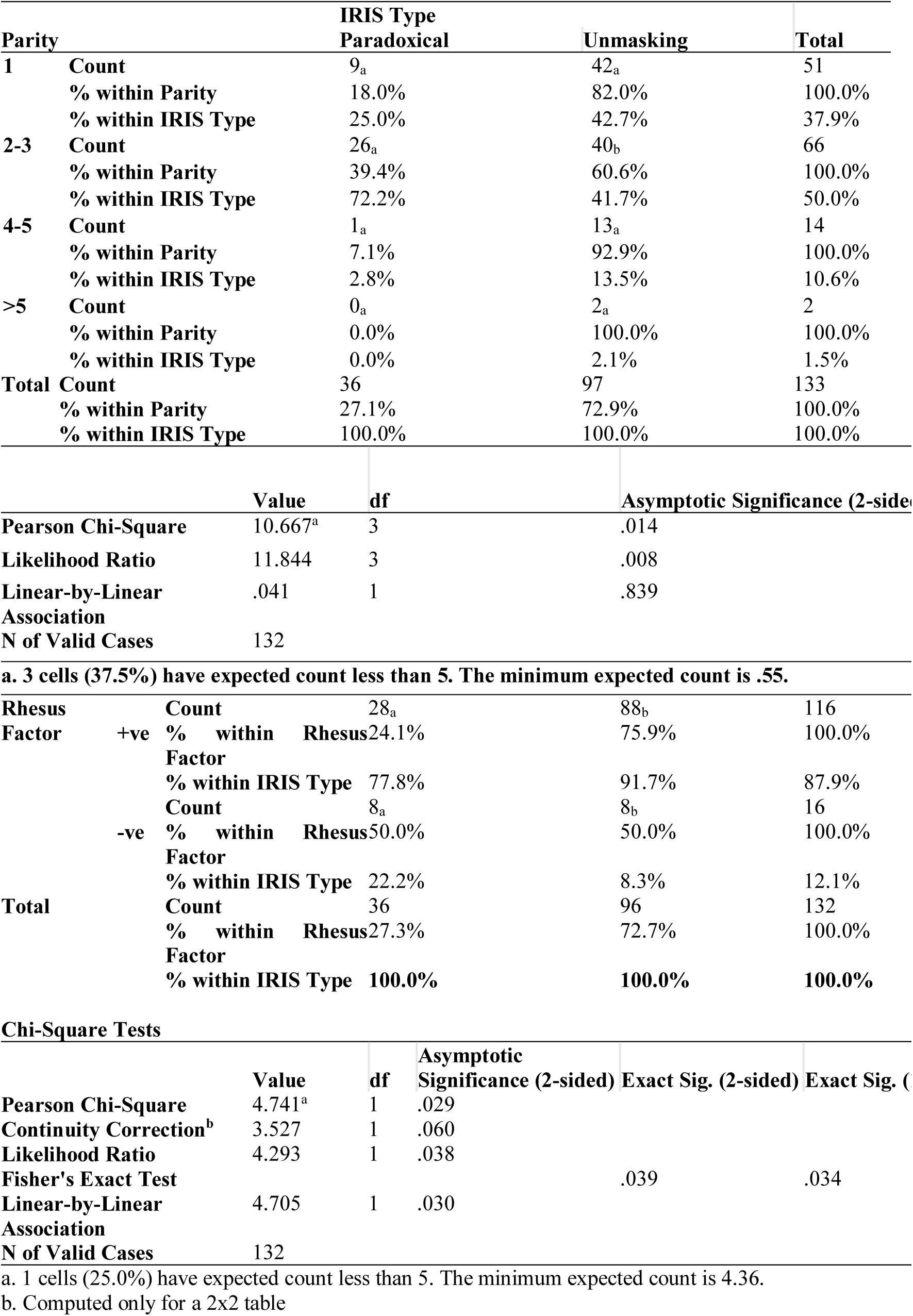
Paradoxical and unmasking IRIS relative to parity and rhesus factor.

### Cumulative incidence of APOs and association with paradoxical and unmasking IRIS

The cumulative incidence of APOs was 20.6% (n = 27) with 55.6 % (n=15) due to unmasking IRIS as compared to 44.4% (n=12) of paradoxical IRIS. Analysis showed a statistically significant association between the IRIS type and the occurrence of an APO χ (1) = 4.911^a^, *P* = 0 .027, demonstrating that, unmasking and paradoxical IRIS differently influence the occurrence of an APO. Further to this, Phi and Cramer’s V tests demonstrated that, the strength of association was very strong (Phi = -191, Cramer’s V = 191, P= 0.027); *table 3*.

**Table 3:**
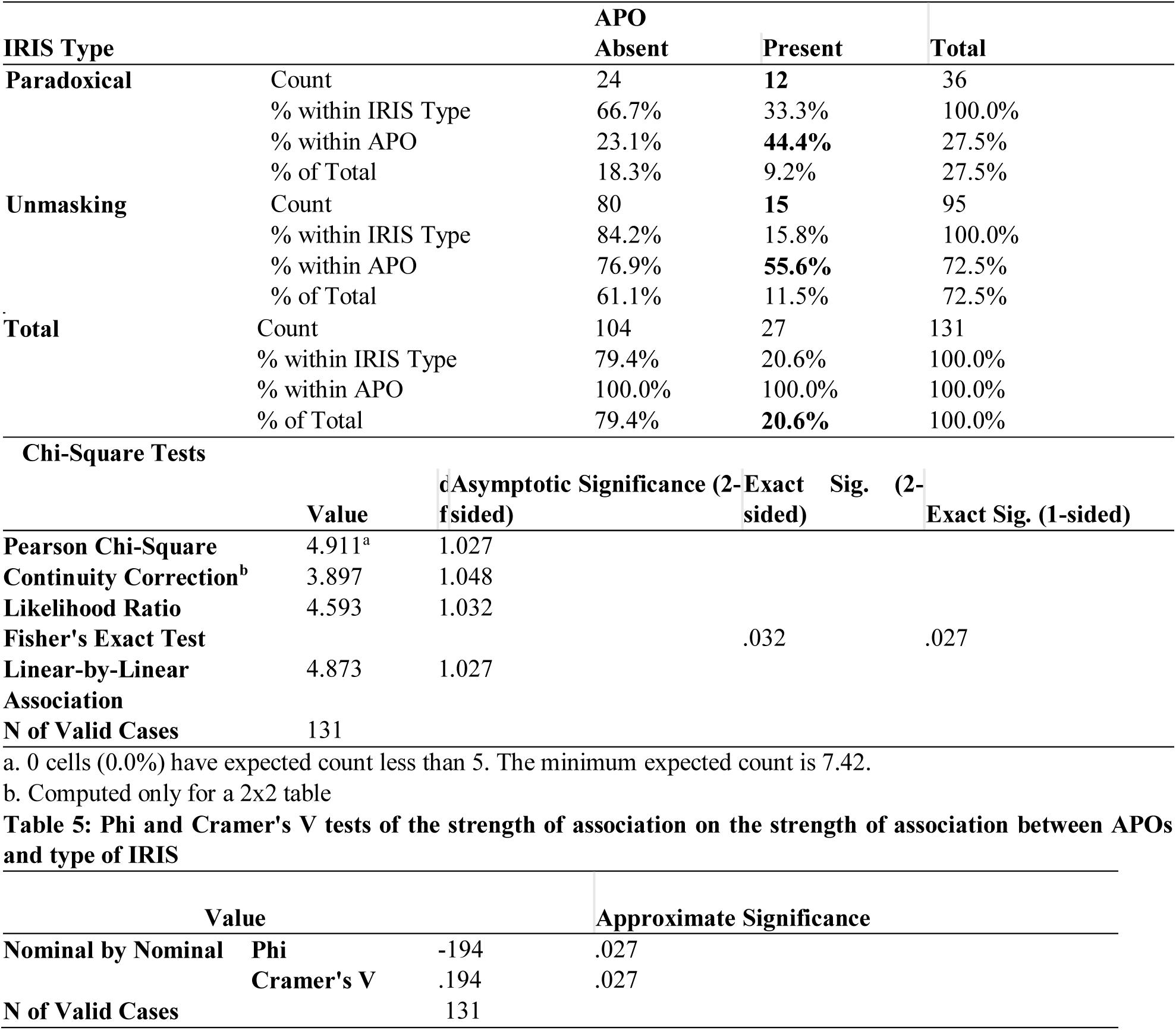
Association between the IRIS type and the occurrence of an APO.

### Multivariate Cox-regression analysis for Pre-ART demographic indicators of paradoxical and unmasking IRIS incidence

Demographic characteristics included in this analysis showed that; pregnant women within the maternal age group (40-49 years) had a positive coefficient towards unmasking IRIS as opposed to paradoxical IRIS, however, this observation was statistically insignificant [(β)=0.329, Wald test (β^) = 1.011, (HR = 1.389, 95% C.I 0.732 - 2.638, P = 0.325]. Further, the occupation status on the contrary demonstrated highly statistically significant coefficients with paradoxical IRIS (protective effect from unmasking IRIS); civil servant [(β)=-3.769, Wald test (β^) = 9.516, (HR = 0.023, 95% C.I 0.002 – 0.253, P = 0.002], self-employed [(β)=-3.769, Wald test (β^) = 9.516, (HR = 0.012, 95% C.I 0.001 – 0.147, P = 0.001]. This was seen with marital status where being married, windowed and or separated were all protective against unmasking IRIS, predicting paradoxical IRIS all with a negative (β) coefficient and a significant P value (< 0.05); *Table 4*.

**Table 4:** Cox-regression analysis for Pre-ART demographic indicators.

|  |  |  |  |  |  |  | 95.0% CI for Exp(B) |  |
| --- | --- | --- | --- | --- | --- | --- | --- | --- |
| Variable | B | SE | Wald | df | Sig. | Exp(B) | Lower | Upper |
| **Maternal Age:** |  |  |  |  |  |  |  |  |
| (20-29) |  |  | 1.011 | 2 | .603 |  |  |  |
| (30-39) | .111 | .241 | .210 | 1 | .647 | 1.117 | .696 | 1.793 |
| (40-49) | .329 | .327 | 1.011 | 1 | .315 | 1.389 | .732 | 2.638 |
| **Education:** |  |  |  |  |  |  |  |  |
| (No formal education |  |  | 4.594 | 3 | .204 |  |  |  |
| (Primary) | -.023 | .457 | .003 | 1 | .960 | .977 | .399 | 2.393 |
| (Secondary) | -.577 | .362 | 2.531 | 1 | .112 | .562 | .276 | 1.143 |
| (Higher/University) | -.230 | .455 | .255 | 1 | .613 | .794 | .325 | 1.940 |
| **Occupation:** |  |  |  |  |  |  |  |  |
| (Unemployed) |  |  | 12.174 | 3 | .007 |  |  |  |
| (Civil servant) | -3.769 | 1.222 | 9.516 | 1 | .002 | .023 | .002 | .253 |
| (Self-employed) | -4.401 | 1.266 | 12.078 | 1 | .001 | .012 | .001 | .147 |
| (Other) | -3.789 | 1.196 | 10.038 | 1 | .002 | .023 | .002 | .236 |
| Religion | -.460 | .573 | .645 | 1 | .422 | .631 | .205 | 1.940 |
| **Marital status:** |  |  |  |  |  |  |  |  |
| (Single) |  |  | 9.100 | 4 | .059 |  |  |  |
| (Married) | -3.207 | 1.111 | 8.332 | 1 | .004 | .040 | .005 | .357 |
| (Separated) | -3.271 | 1.104 | 8.773 | 1 | .003 | .038 | .004 | .331 |
| (Windowed) | -2.943 | 1.185 | 6.171 | 1 | .013 | .053 | .005 | .537 |
| (Not recorded) | -3.657 | 1.524 | 5.762 | 1 | .016 | .026 | .001 | .511 |

### Survival and hazard function at the mean of demographic covariates

Generally, the cumulative survival function evaluating all the demographic covariates at their mean showed that, post ART initiation, over 80% of the pregnant women survived before IRIS diagnosis up to about six weeks while half (0.5) of the entire population of pregnant women had been diagnosed with IRIS at approximately 8.5 weeks, after which survival ability before IRIS diagnosis decreased marginally towards zero; *figure 2*.

**Figure 2:**
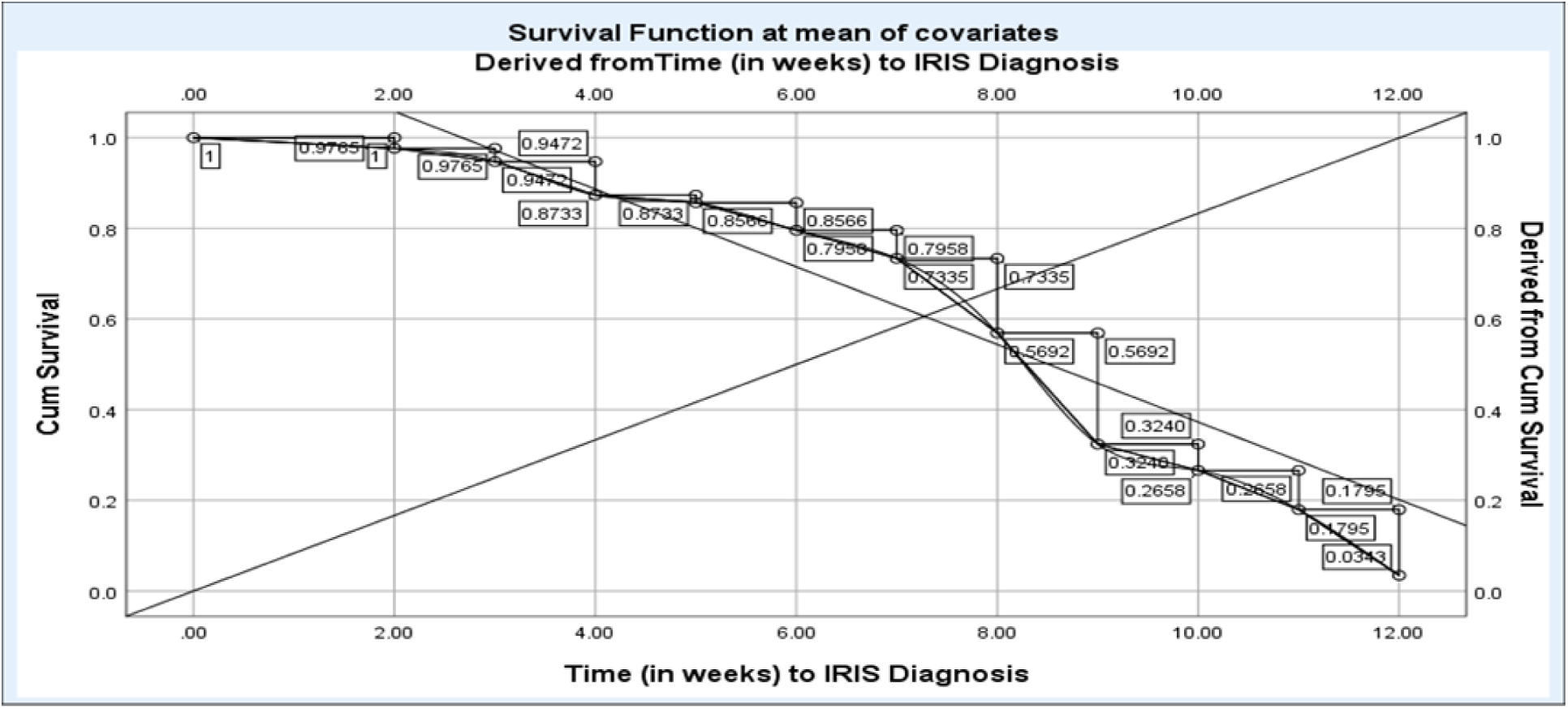
Survival function at the mean of demographic covariates.

Notably, the hazard (IRIS diagnosis) increased with the increasing age where by, pregnant women between 20-29 years experienced the IRIS diagnosis at a lower rate as compared to older women (30-39 years) and (40-49 years), a differential aspect that was more feasible and clearer from around week 7 post ART initiation; *figure 3*.

**Figure 3:**
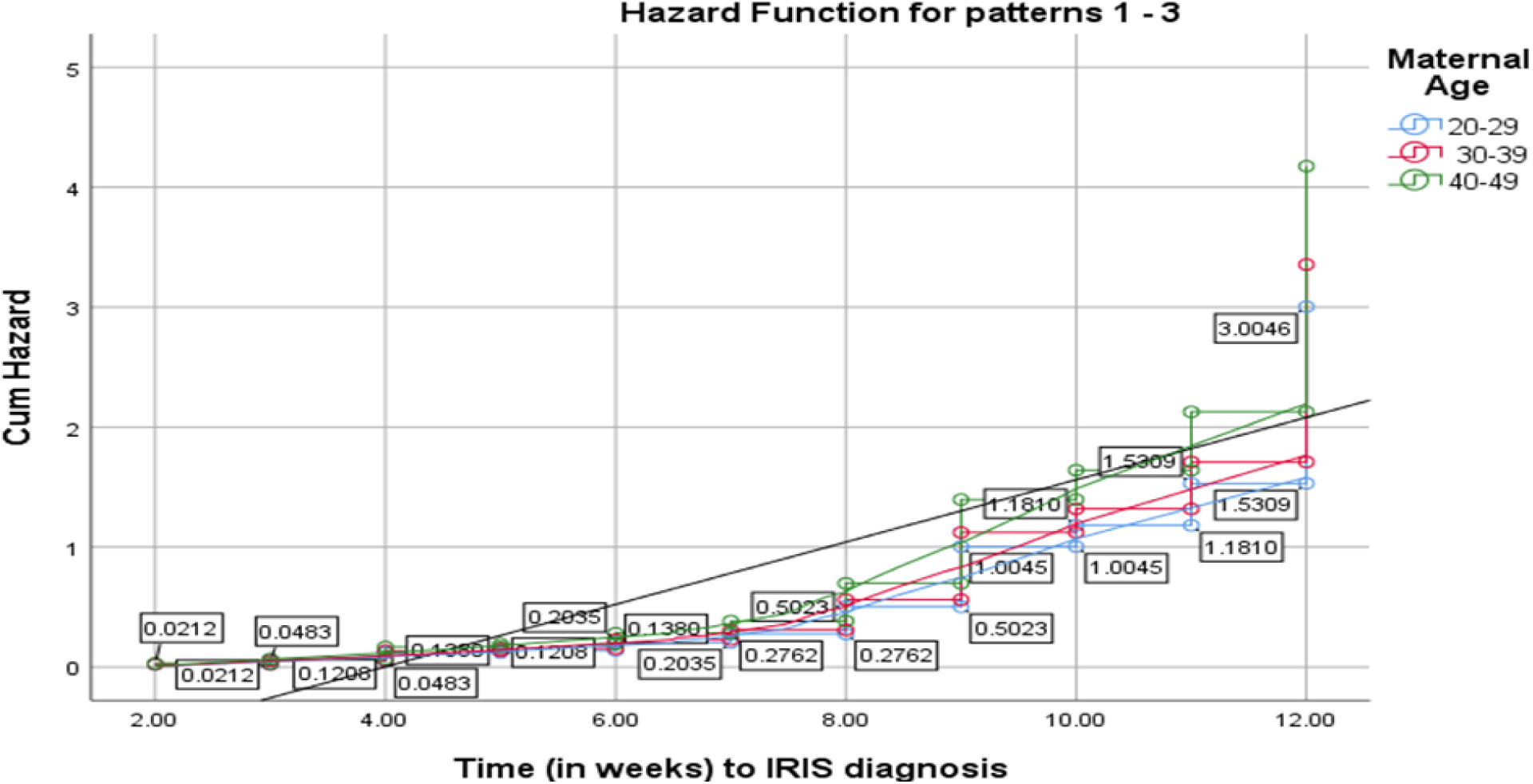
Hazard function for IRIS diagnosis by age.

**Figure 4:**
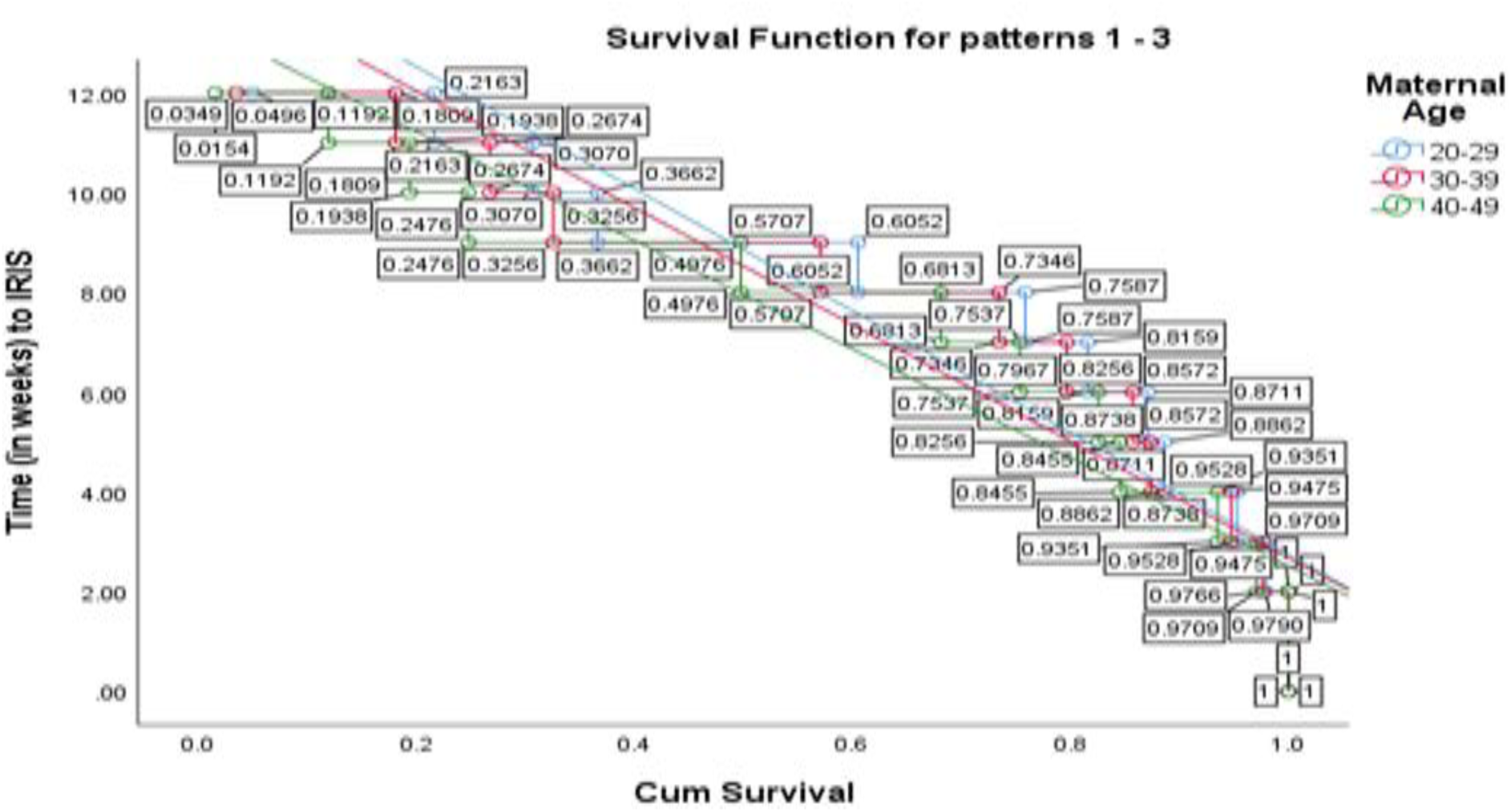
Survival function for IRIS diagnosis by age.

### Kaplan Meier Survival estimates of APOs by the type of IRIS

The survival probability as a function of time was obtained to ascertain the survival to APO by either paradoxical or unmasking IRIS. Kaplan-Meier survival curves for the cumulative risk of APO among ART naïve pregnant women who developed paradoxical IRIS compared with ART naïve pregnant women who developed unmasking IRIS showed that, women presenting with unmasking IRIS appeared to have a higher survival rate than the women developing paradoxical IRIS (χ2 = 5.292, Log Rank test = 0.021); *table 5*.

**Table 5:**
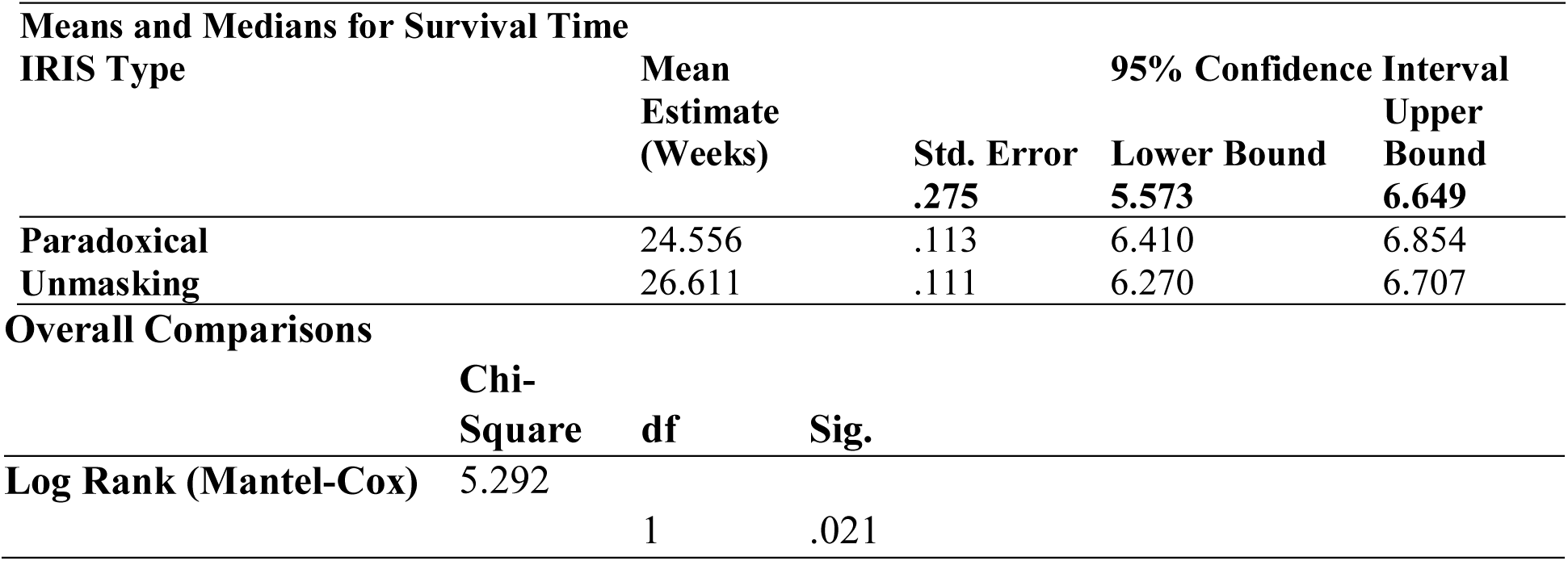
Kaplan Meier Survival estimates of APOs by the type of IRIS.

Within the first to second month post IRIS identification, APOs were seemingly not experienced from both groups, however, from second month onwards, APOs were experienced at a different rate with over 80% (unmasking) and about 66% (paradoxical) surviving APOs at the end of 28^th^ week (7 months) post IRIS identification, that is, at the end of the third trimester. The median survival time for women diagnosed with paradoxical and those diagnosed with unmasking IRIS was the same; *figure 10.* Consequently, both unmasking and paradoxical IRIS groups had similar cumulative hazard of APOs up to around 2 months from where, the differences started showing clearly with paradoxical IRIS demonstrating a higher cumulative hazard rate for APOs as compared to unmasking ending at HR = 0.4 and 0.18, respectively; *figure 11*

**Figure 10:**
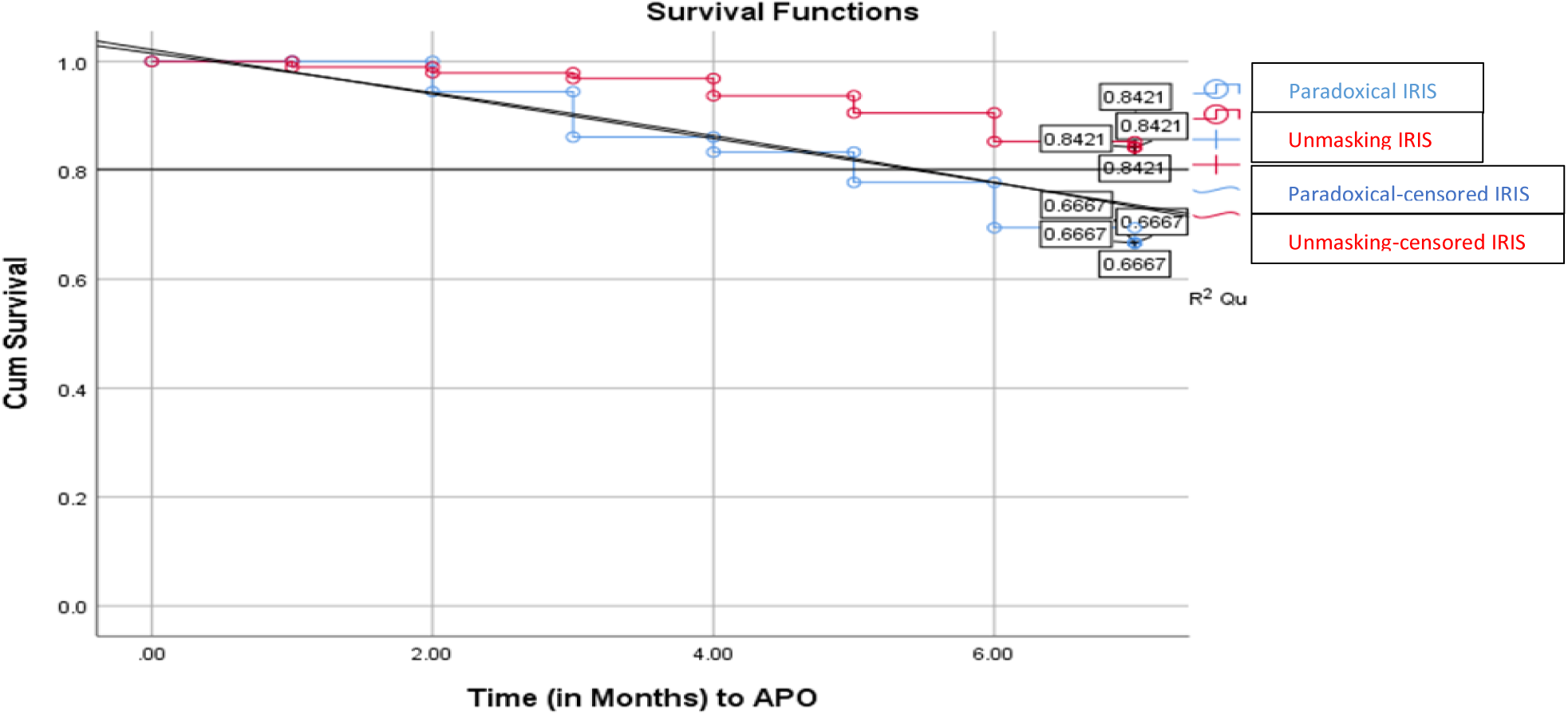
Survival plot Kaplan Meir curve on APO between unmasking and paradoxical IRIS

**Figure 11:**
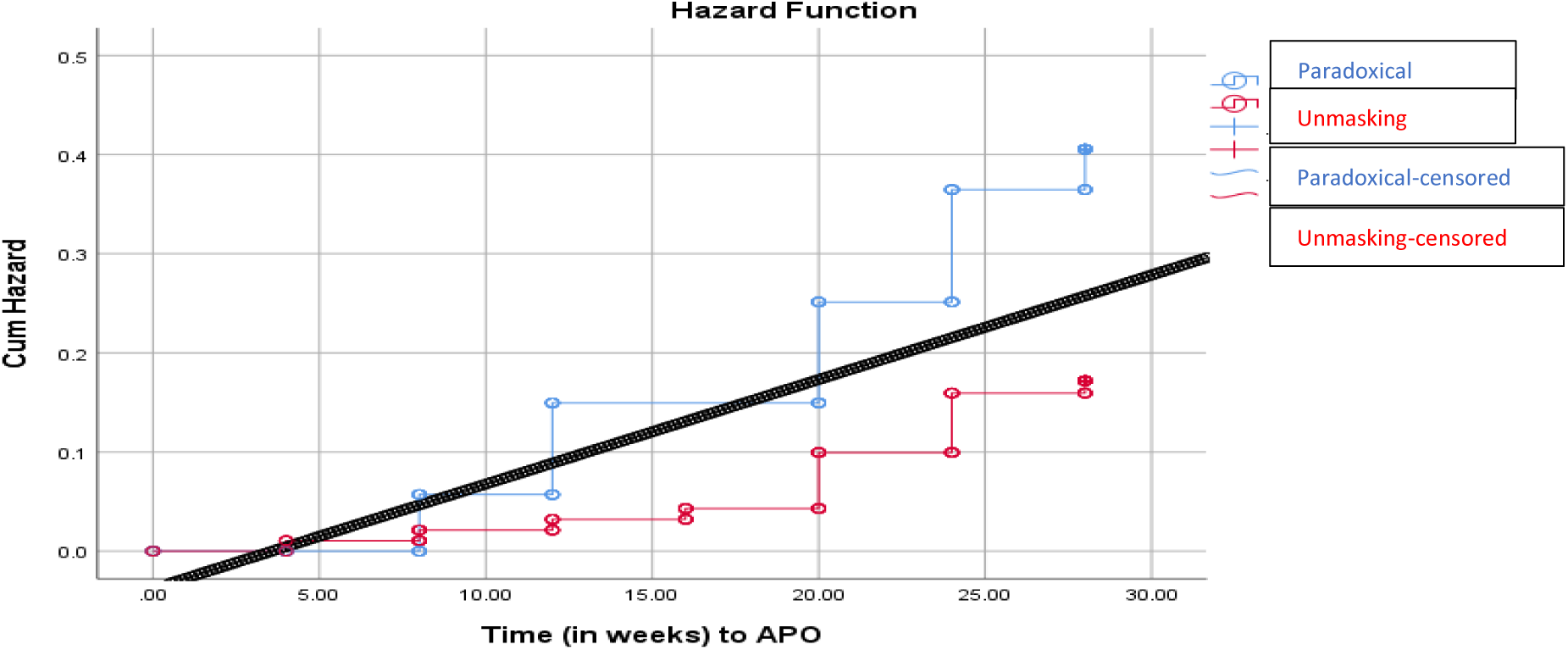
Hazard plot based on hazard function Kaplan Meir curve on APO between unmasking and paradoxical IRIS

Collectively, for exploratory and confirmatory classification analysis to assess their predictions of APOs after IRIS identification, decision tree analysis was performed. This established that, 25% of the 69 women aged 30-39 years experienced APOs as compared to 16% of the 62 women aged 20-29 and 40-49 years combined (P = 0.688); *table 6.* Similarly, an APO was predicted close to the perfect prediction model; *figure 12*. The ratio of the node response percentage for the target category compared to the overall target category response percentage for the entire sample; *figure 13*.

**Figure 12.**
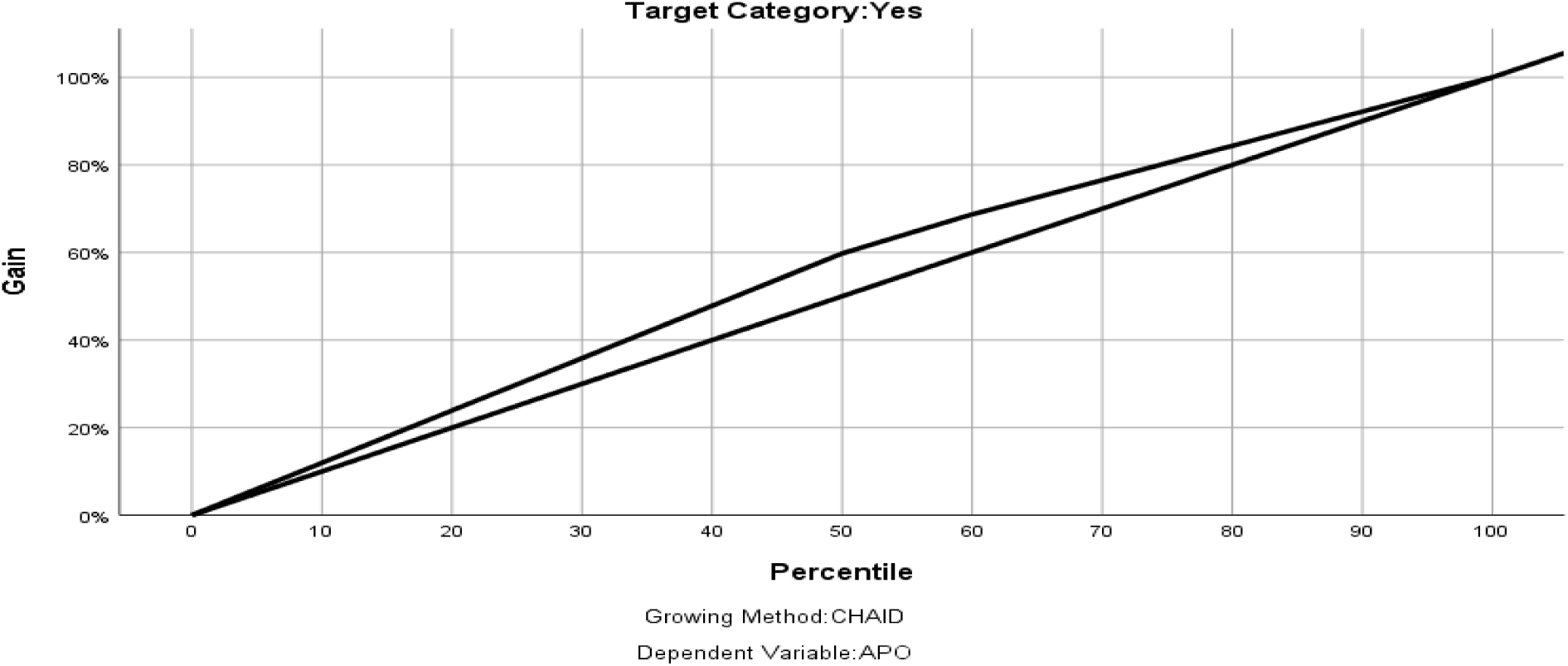
The gain chart curve on the prediction model of APOs

**Figure 13.**
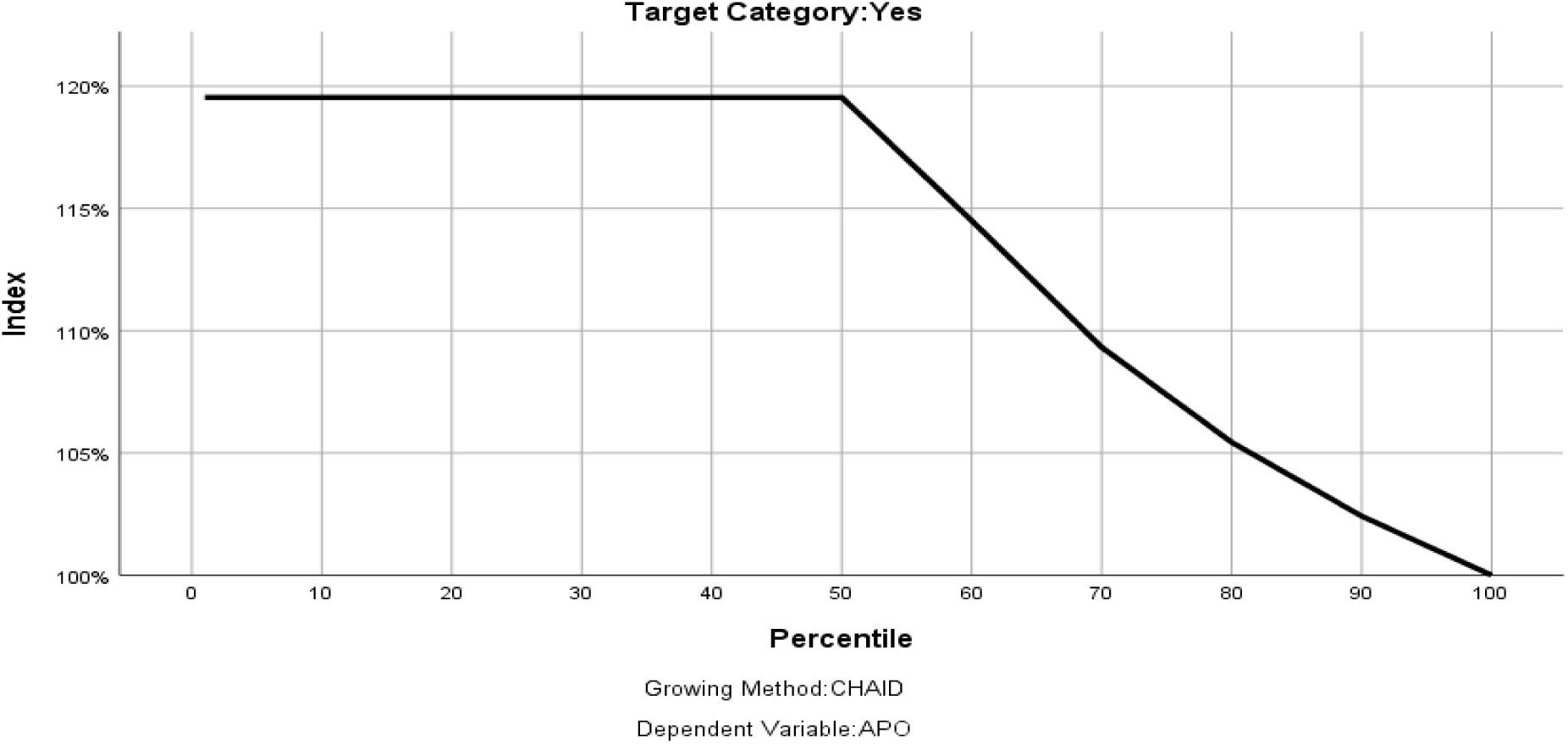
The index chart shows the ratio of the node response percentage

**Table 6:** Decision tree analysis table demonstrating specific demographics relative to IRIS occurrence.

| Node | No-APO |  | APO |  | Total |  | Predicted Category | Parent Node | Primary Independent Variable |  |  |  |  |
| --- | --- | --- | --- | --- | --- | --- | --- | --- | --- | --- | --- | --- | --- |
|  | N | Percent | N | Percent | N | Percent |  |  | Variable | Sig.<sup>a</sup> | Chi-Square | df | Split Values |
| 0 | 104 | 79.4% | 27 | 20.6% | 131 | 100.0% | No |  |  |  |  |  |  |
| 1 | 52 | 83.9% | 10 | 16.1% | 62 | 47.3% | No | 0 | Maternal Age | .688 | 1.445 | 1 | 20-29; 40-49 |
| 2 | 52 | 75.4% | 17 | 24.6% | 69 | 52.7% | No | 0 | Maternal Age | .688 | 1.445 | 1 | 30-39 |

Growing Method: CHAID
Dependent Variable: APO
a. Bonferroni adjusted

## Discussion

In this active records-based prospective cohort study, we evaluated IRIS incidence, demographic indicators, survival-time to and prediction of APOs using the Cox model.

The estimated prevalence of HIV-IRIS from this current findings was consistent with studies reporting that, up to 25% to 30% of HIV patients who are on antiretroviral therapy developed IRIS[21], [22]. Majorly, HIV infected Pregnant women with a parity of one were diagnosed with paradoxical IRIS, where in general, parity of a woman was linked with a specific type of IRIS. This has not been clearly ascertained by other studies however, closer finding tends to demonstrate such direction[23], [24].A positive rhesus factor screening in the first trimester was associated with unmasking IRIS. This finding though not out rightly identified before, it can seemingly be established under some studies[25], [26].

The cumulative incidence of APOs was 20.6% among ART naïve pregnant women, a very precise similarity with another study that showed a total of 149 singleton pregnancies had documented APOs at 116 (20.6%) in HIV-positive pregnancies[27]. Closer findings have also been feasible with other studies[17], [28]. With higher rates due to unmasking IRIS as compared paradoxical IRIS, this has been shown by some studies though not fully definitive[9], [29]. Notably, difference opportunistic infections and clinical presentations accounted to specific type of IRIS although not clearly demonstrated among some studies[30], [31].

Women who were pregnant at an older maternal age group (40-49 years) demonstrated an inclination towards unmasking IRIS as opposed to paradoxical IRIS. Although not a clear evidence, Ireland findings tend to explain the current study’s outcome[32]. Occupation status in reference to being a civil service clearly predicted paradoxical IRIS as it has also been depicted in a study however indirectly, that established employment is positively associated with movement along the HIV continuum of care and that, determinants of employment among PLWH are highly complex[33]. Marital status where being married, windowed and or separated were all predictive of paradoxical IRIS. Subject to this, paradoxical IRIS has been found to be more common than unmasking IRIS [34], [35].

After starting ART in pregnant women, majority stayed free of IRIS diagnosis at about one and half months. Clearly, an existing primary study findings have been supported by the current results that, IRIS occurred after about weeks of ART initiation[36]. To implicate this in existing literature, a pool of older studies through a systematic review and meta-analysis have seemingly proved this fact[37]. Additionally, most recent primary studies tend to support this current findings[38], [39], [40].

IRIS diagnosis was found to increase with increasing age as opposed to younger age in our findings similar to a Ugandan based research clearly showing older adults had a poorer immunological response[41]. This is further backed up by a much older study that, that older adults, particularly those aged 50 years and above, are at a higher risk of developing IRIS due to their weakened immune systems and higher likelihood of having underlying opportunistic infections[36].

On cumulative APO by either paradoxical or unmasking IRIS, ART naïve pregnant women diagnosed with unmasking IRIS appeared to have longer survival rate than those diagnosed with paradoxical IRIS before experiencing any form of an APO. This concurs indirectly with another study’s finding that, paradoxical IRIS is associated with a higher risk of adverse pregnancy-fetal outcomes (APOs) such as preterm birth, low birth weight, and stillbirth[42]. On the flipside, unmasking IRIS has been found to be milder relative to APOs as similarly found in another study that, women diagnosed with unmasking IRIS tend to have a longer survival rate and better pregnancy outcomes compared to those with paradoxical IRIS[43].

Older pregnant women were seemingly diagnosed with unmasking IRIS as opposed to paradoxical IRIS in the current study. This finding is supported by existing research that established older adults had four times greater odds (OR = 4.7 (3.1–7.0)) of having an HIV comorbidity compared to younger adults and specifically closer to the current study population, being female (OR = 1.6 (1.1–2.4) increased the odds of HIV comorbidity[44]. This could be attributed to factors such as a weakened immune system and a higher likelihood of having preexisting opportunistic infections [45], [46], [47]. Further, self-employed and civil servant pregnant ART naïve women were highly diagnosed with paradoxical IRIS and however having no clear existing findings similar to this factors such as baseline CD4 count, age, and the presence of opportunistic infections are known to influence the risk of developing IRIS. Notably, being married, windowed and or separated predicted paradoxical IRIS as opposed to unmasking IRIS.

After ART initiation, over 80% of the pregnant women were not diagnosed with HIV-IRIS until up to about six weeks, a consistent finding with an existing information[48], [49] while half (0.5) of the entire IRIS population of pregnant women had been diagnosed with IRIS at precisely over a period of two-months, a fact that is implicated by another study[50]. Notably, IRIS diagnosis increased with the increasing age as supported earlier studies[51]. Further, in general, older age may be associated with decreased immunity that may be a clear predictor of quicker diagnoses with IRIS as opposed to younger age [52].

ART naïve pregnant women diagnosed with unmasking IRIS appeared to have a higher survival rate than the women developing paradoxical IRIS in our current finding. This has been mimicked by another much older study as well[53], and implied in more recent findings[54], [55], [56], [57], [58]. In regards to the incidence of APOs relative to the type of IRIS, women with paradoxical IRIS seemingly experienced APOs much earlier than those who had unmasking IRIS. This has been demonstrated by another study although not explicitly focusing on pregnant women[59].

Decision tree analysis demonstrated that, ART naïve pregnant women aged 30 to 39 years experienced majority of APOs as compared to much younger and much older ART naïve pregnant women. This observation though did not portray a significant association. It is of course expected that, women over 40 years are at more risk for APOs [60], [61], [62]. However, finding is supported by the fact that, fewer women would conceive at over 40 years as compared to at 30 years. This is by noting that women between 30 and 39 years would have most pregnancies and as such, proportion wise, they may encounter a similar trajectory of APOs demonstrated in the current results.

This study had some limitations. First, the APOs identification was cumulative without ascertaining a specific APO due to specific demographic relative to IRIS occurrence; however, the main purpose for the study was establish just a possible prediction of APO irrespective of the type. Again, the survival time to an APO wasn’t based on a specific type but in the context of the study, the hazard risk (APO) was treated as such despite its nature. Last, ages at pregnancy were diverse of which, IRIS incidence, survival time to APO and the related demographics would different influence the APOs. This however was mitigated by decision tree analysis to evade such a possible doubt. Despite these limitations, the study provides feasible information about the incidence of IRIS by type, whether unmasking or paradoxical. These results estimate the survival time to an APO while accounting possible predictors of APOs ART-naïve pregnant women relative to IRIS.

## Conclusions

About a quarter of ART naïve pregnant women experienced IRIS by the 12^th^ week post ART initiation and IRIS diagnosis risk increased with older maternal age group which also, mostly predicted unmasking IRIS. Majorly, women with a parity of one and a positive rhesus factor screening experienced unmasking IRIS with those of 2-3 parity, experiencing paradoxical IRIS. Most APOs were predicted by unmasking IRIS however with a higher survival rate than the women developing paradoxical IRIS who demonstrated a higher cumulative hazard rate for APOs. Most APOs were among women aged 30-39 years.

## Data Availability

All data produced in the present study are available upon reasonable request to the authors

## Notes

### Competing Interest Statement

The authors have declared no competing interest.

### Funding Statement

This study was partially funded by Kenya Medical Training College

### Author Declarations

Ethics committee of Kenyatta National Hospital and University of Nairobi gave ethical approval for this work

